# Risk factors for fecal carriage of multidrug-resistant *Escherichia coli* in a college community: a penalized regression model

**DOI:** 10.1101/2020.11.16.20231969

**Authors:** Yuan Hu, Julia Rubin, Kaitlyn Mussio, Lee W. Riley

## Abstract

**Background:** Bacterial antimicrobial resistance is a serious global public health threat. Intestinal commensal drug-resistant bacteria have been suggested as an important reservoir of antimicrobial resistant genes (ARGs), which may be acquired via food. We aimed to identify risk factors associated with fecal carriage of drug-resistant commensal *Escherichia coli (E. coli)* among healthy adults focused on their dietary habit.

**Methods:** We conducted a cross-sectional study targeting healthy adult volunteers in a college community. Fecal samples and questionnaires were obtained from 113 volunteers. We conducted backward elimination logistic regression and least absolute shrinkage and selection (LASSO) methods to identify risk factors.

**Results:** We analyzed responses from 81 of 113 volunteers who completed the questionnaire. The logistic regression and LASSO methods identified red meat consumption to be associated with increased risk (OR = 6.13 [1.83-24.2] and 1.82, respectively) and fish consumption with reduced risk (OR = 0.27 [0.08-0.85] and 0.82) for the carriage of multidrug-resistant *E. coli*, adjusted for gender, employment status, frequently-used supermarket, and previous travel.

**Conclusions:** Dietary habits are associated with the risk of fecal carriage of multidrug-resistant *E. coli*. This study supports the growing evidence that food may be an important source of ARGs present in human commensal *E. coli*.

## 1. Introduction

Antimicrobial drug resistance is one of the most pressing global public health concerns of our time, threatening the effective prevention and treatment of infectious diseases in every country [1] [2]. In the United States, U.S. Department of Health & Human Services have declared the necessity to take national action and to strengthen surveillance system to combat the spread of antimicrobial resistance [3]. In particular, several drug-resistant Gram-negative bacterial species belonging to Enterobacteriaceae have come to be designated by Centers for Disease Control and Prevention (CDC) and the World Health Organization (WHO) to be “urgent-threat” or “priority 1” pathogens [4].

Among the Gram-negative bacterial pathogens, *Escherichia coli* (*E. coli*) is the most frequent cause of common extraintestinal infections, including urinary tract infections (UTI) and bloodstream infections (BSI) [5]. They are referred to as extraintestinal pathogenic *E. coli* (ExPEC) [6]. The prevalence and incidence of infections caused by drug-resistant ExPEC have been rapidly increasing worldwide [7,8].

Major sources of drug-resistant Gram-negative bacteria include the environment such as contaminated water [9], food including meat [10,11] and vegetables [12,13], and healthcare settings [14]. Additionally, intestinal commensal drug-resistant bacteria have been reported as an important reservoir of antimicrobial drug resistance genes (ARGs) [15,16]. *E. coli* is a member of the commensal flora of human and other warm-blooded animal intestinal tracts. As such, they can acquire ARGs by horizontal gene transfer [17] from drug-resistant *E. coli* strains and other Gram-negative bacteria that enter the intestinal tract via exposures to contaminated food, water, and other external sources. *E. coli* can be transmitted through contaminated water or food, or through contact with people and other animals [18]. Thus, risk factors for fecal carriage of drug-resistant commensal *E. coli* and ARGs could include exposures to environmental sources of drug-resistant bacteria, in addition to traditional risks such as prior use of antibiotics [19,20] or healthcare-associated infections [21,22].

In our previous study, we found 81% of healthy volunteers at a college community to harbor intestinal drug-resistant commensal Gram-negative bacteria [23]. They included 12 species of Gram-negative bacteria that harbored a variety of ARGs and integrons that are frequently found among extraintestinal pathogenic *E. coli* (ExPEC) [23]. Risk factors for the carriage of these ARGs, however, were not determined.

We first conducted a systematic review and meta-analysis on risk factors of intestinal carriage of drug resistant *E. coli* [24], but found no studies from North America that met the criteria to be included in the review, even though North America is a major food-exporting region in which antibiotics are heavily used in food animal husbandry and agriculture. Also, we could not find studies that particularly focused on dietary behavior as a risk factor for intestinal carriage of drug-resistant *E. coli* or ARGs. Two studies reported significant association between food and carriage of drug resistant bacteria. One study reported that raw milk consumption increases the risk of intestinal colonization of drug resistant bacteria [25], and another reported food consumption from street vendors as a travel-associated risk factor [26].

Here, we designed and implemented a cross sectional study to identify dietary risk factors associated with fecal carriage of multidrug-resistant commensal *E. coli* among healthy adult population in a college community. We focused on the carriage of multidrug-resistant *E. coli* in particular because of its clinical relevance to diseases such as UTI and BSI. Because of the relatively small sample size of the study, we used multiple logistic regression and penalized regression methods to select the best model to identify risks.

## 2. Materials and Methods

### 2.1 Sample collection, antimicrobial susceptibility screening, and questionnaires

As previously described by Rubin et al. [23], we prospectively collected and cultured fecal swab samples from 113 healthy volunteers at a university campus in northern California between June and October 2018. Eligible participants included both men and women between 18 and 65 years of age, with no medical history of urinary tract corrective surgery or abnormality, or bladder catheterizing or hospitalization within the 6 months prior to sample collection. At recruitment, participants were provided a collection kit containing a Blair transport media rectal swab (Becton Dickinson BBL), two bio-safety bags, and detailed collection instructions. Each kit also included a questionnaire regarding antibiotic use, history of UTI, as well as diet and lifestyle characteristics. Participants were instructed to send the swab and the completed questionnaire back to the laboratory via USPS mail immediately after collection. Once delivered, the study coordinator analyzed the fecal swab samples within 48 hours. Detailed microbiologic procedures, including antimicrobial drug susceptibility testing of the *E. coli* isolates, are described in the report by Rubin et al [23].

### 2.2 Survey

The survey questions focused on dietary and behavioral habits that might be associated with the intestinal acquisition of ARG. We asked participants to recall their habits within the 1-year period before completion of the survey questionnaire. The survey instrument was devised partly based on the standard questionnaire used by the Centers for Disease Control and Prevention, Foodborne Disease Outbreak and Surveillance Unit, National Hypothesis Generating Questionnaire (https://www.cdc.gov/foodsafety/outbreaks/surveillance-reporting/investigation-toolkit.html) and was partly based on the instrument used by Manges et al. in a 2007 study investigating the links between retail meat and risk of UTI [27]. Participants were asked to respond according to the following scale: never, past day (24 hours), past week, past month, past year, or more than a year ago. Responses to these questions were dichotomized to reflect modest versus frequent behavior, where the response category that corresponded to a behavior within the past month was defined as frequent. The following information was requested of the participants: biological sex, academic status, meat consumption, dietary restrictions, dairy and egg consumption, raw meat consumption, raw vegetable consumption, preference of organically produced food, number of live-in housemates, location of meal preparation, possession of companion animals, number of sexual partners, history of antibiotic use, history of hospitalization, history of UTI, placement and type of a intrauterine device, and location and duration of travel outside of the United States.

### 2.3 Dataset construction

To identify the risk factors associated with intestinal carriage of antimicrobial resistance, we compiled a set of 66 variables from our survey that detailed their diet habit, lifestyle, and past antibiotic use. All questions were dichotomized based on frequent (“Yes”) or modest (“No”) response as described above. We excluded variables for which (1) less than 5% or more than 95% of the participants replied “Yes” and (2) more than 10% of the participants answered “Do not know” or NA. We merged the results from antimicrobial resistance testing to the survey results. Relationships between each set of variables were assessed with correlation coefficients. In this study, we defined isolates with resistance to two or more antimicrobial drug classes as multidrug-resistant and those with resistance to only one drug as drug-resistant.

### 2.3 Statistical analysis

All statistical analyses were conducted with R version 3.5.1 [28]. First, univariate and multivariate logistic regression analysis was performed. In the multivariate analysis, a model with all potential risk factors were constructed. Then we used R package ‘stats’ (version 3.5.1) ‘step’ function with a backward elimination method to determine the best-fitting model. The best-fitting model was determined with Akaike information criteria (AIC) [29]. All tests performed were two-sided, and a P-value <0.05 was considered significant.

Penalized regression with the least absolute shrinkage and selection operator (LASSO) models [30] were used to identify variables associated with carriage of multidrug-resistant *E. coli*. Association with drug resistance was not tested with LASSO regression due to the high imbalance between the prevalence of drug resistant *E. coli* and drug susceptible *E. coli*. The LASSO regression functioned as a variable selection process, which reduced the variables to a subset of variables that were consistently related to multidrug resistance. We conducted 10-fold cross validation to select the best fitting model with the minimum mean squared error (MSE). Standard errors were not assessed for LASSO regression model because of its strong bias arising from penalized estimation method. All LASSO regression was performed with R package ‘glmnet’ (version 2.0.18).

The model performance was evaluated by receiver operating characteristic (ROC) curves and the area under the curve (AUC) was used to classify the participants with and without multidrug-resistant *E. coli*. Hosmer-Lemeshow fit test was used to assess the agreement between observed and model-predicted proportions of carriage of multidrug-resistant *E. coli* [31]. The difference of AUCs was tested by a nonparametric approach developed by DeLong et al [32].

## 3. Results

### 3.1 Background information

Between June and October 2018, 113 fecal swab samples were collected and cultured, and Gram-negative bacteria were isolated from each culture. Of 113 volunteers, 103 returned both stool sample and the questionnaire. Of 103 stool samples, 93 yielded *E. coli* colonies on MacConkey plates. Of these, 81 corresponding survey questionnaires had complete response without unanswered questions (Figure 1).

**Figure 1:**
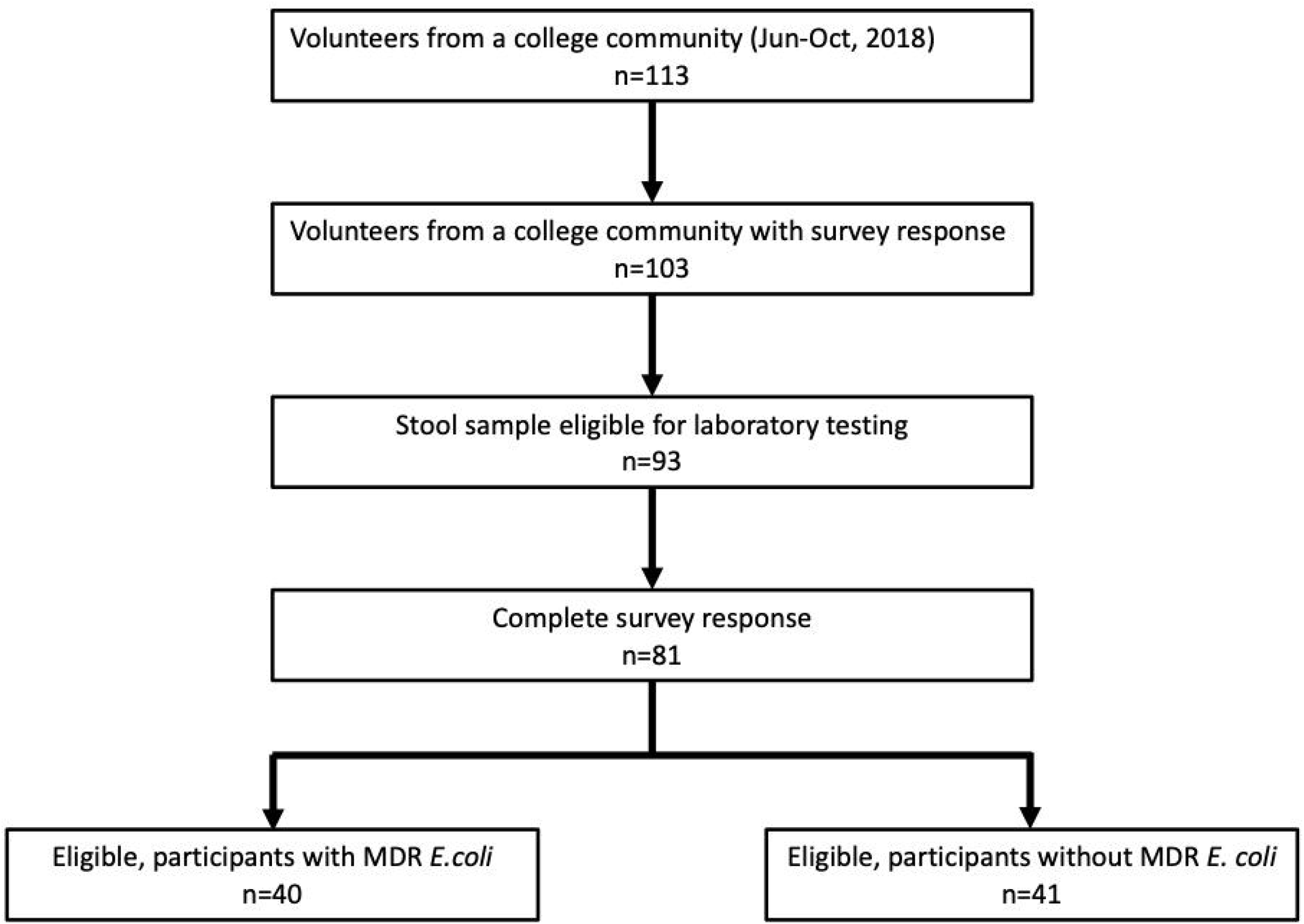
Flowchart of study samples

The participants were adults between 18 to 65 years of age (Table 1). Seventy-eight (84%) of 93 participants whose stool sample was analyzed had *E. coli* resistant to either ampicillin, trimethoprim-sulfamethoxazole, gentamicin, or colistin, each representing a different class of antimicrobial agent. Of these, 48 (52%) were multidrug resistant, defined as resistance to two or more classes of antimicrobial agents, 30 (32%) were resistant to only one drug class. Fifteen (16%) were susceptible to all the tested drugs.

**Table 1:** Univariate analysis of potential risk factors for fecal carriage of MDR *E. coli*

|  | No. of population (%) | No. of samples with MDR *E. coli* (%) | OR [CI95%] |
| --- | --- | --- | --- |
| Gender (Female) | 49 (61) | 29 (59) | 2.77 [1.12-7.17] |
| **Employment** |  |  |  |
| Undergrad student | 19 (24) | 14 (74) | 3.88 [1.31-13.24] |
| Graduate student | 37 (46) | 15 (41) | 0.52 [0.21-1.25] |
| Staff | 23 (28) | 10 (44) | 0.72 [0.27-1.89] |
| **Dietary habit** |  |  |  |
| Poultry | 64 (79) | 34 (53) | 2.08 [0.70-6.67] |
| Red meat | 52 (64) | 30 (58) | 2.59 [1.03-6.85] |
| Fish | 31 (38) | 12 (39) | 0.50 [0.20-1.23] |
| Raw meat/fish | 43 (53) | 22 (51) | 1.16 [0.49-2.81] |
| Raw vegetable | 79 (98) | 39 (49) | 0.98 [0.04-25.24] |
| Diet restriction | 19 (24) | 8 (42) | 0.68 [0.24-1.91] |
| Organic food | 17 (21) | 9 (53) | 1.20 [0.41-3.57] |
| Living with roommate | 71 (88) | 36 (51) | 1.54 [0.41-6.48] |
| Food made at home | 72 (89) | 35 (49) | 0.76 [0.18-3.08] |
| Food from outside home | 30 (37) | 16 (53) | 1.29 [0.52-3.20] |
| **Supermarket** |  |  |  |
| A | 49 (61) | 25 (51) | 1.18 [0.48-2.90] |
| B | 37 (46) | 21 (57) | 1.73 [0.72-4.23] |
| C | 31 (38) | 13 (42) | 0.62 [0.25-1.51] |
| other grocery shop | 31 (38) | 13 (42) | 0.62 [0.25-1.51] |
| **Pet** | 22 (27) | 10 (46) | 0.81 [0.30-2.15] |
| **Travel** |  |  |  |
| travel within one year | 51 (63) | 28 (55) | 1.83 [0.74-4.65] |
| travel longer than 1 month | 27 (33) | 15 (56) | 1.45 [0.57-3.72] |
| Southeast Asia | 4 (5) | 2 (50) | 1.03 [0.12-8.91] |
| East Asia | 10 (12) | 5 (50) | 1.03 [0.27-4.00] |
| Central and South America | 21 (26) | 10 (48) | 0.91 [0.33-2.47] |
| Europe | 27 (33) | 16 (59) | 1.82 [0.72-4.73] |
| **Medical history** |  |  |  |
| previous antibiotics use | 31 (38) | 18 (58) | 1.76 [0.72-4.43] |
| past UTI | 12 (15) | 6 (50) | 1.03 [0.30-3.60] |
| **Others** |  |  |  |
| Living with roommate | 71 (88) | 36 (51) | 1.54 [0.41-6.48] |

We compiled the responses to our questionnaire into a dataset with 66 variables and 103 observations (Supplementary Table 1). Variables with (1) more than 10 “Do not know” or (2) 5% or less “Yes” or “No” were removed from the data to be analyzed. Twenty-six (39%) of 66 variables were eligible for analysis.

Relationships between each of the predictors under consideration are presented in Figure 2 as correlation coefficients matrix. Females were more likely to report past urinary tract infection (UTI). Those reporting previous antibiotics use had strong correlation with previous UTI. We observed a correlation among employment status and travel status. Also, dietary behaviors were correlated among each factor. There were positive correlations among red meat consumption, poultry consumption, but negative correlations among those reported to have dietary restriction such as being vegan or vegetarian.

**Figure 2:**
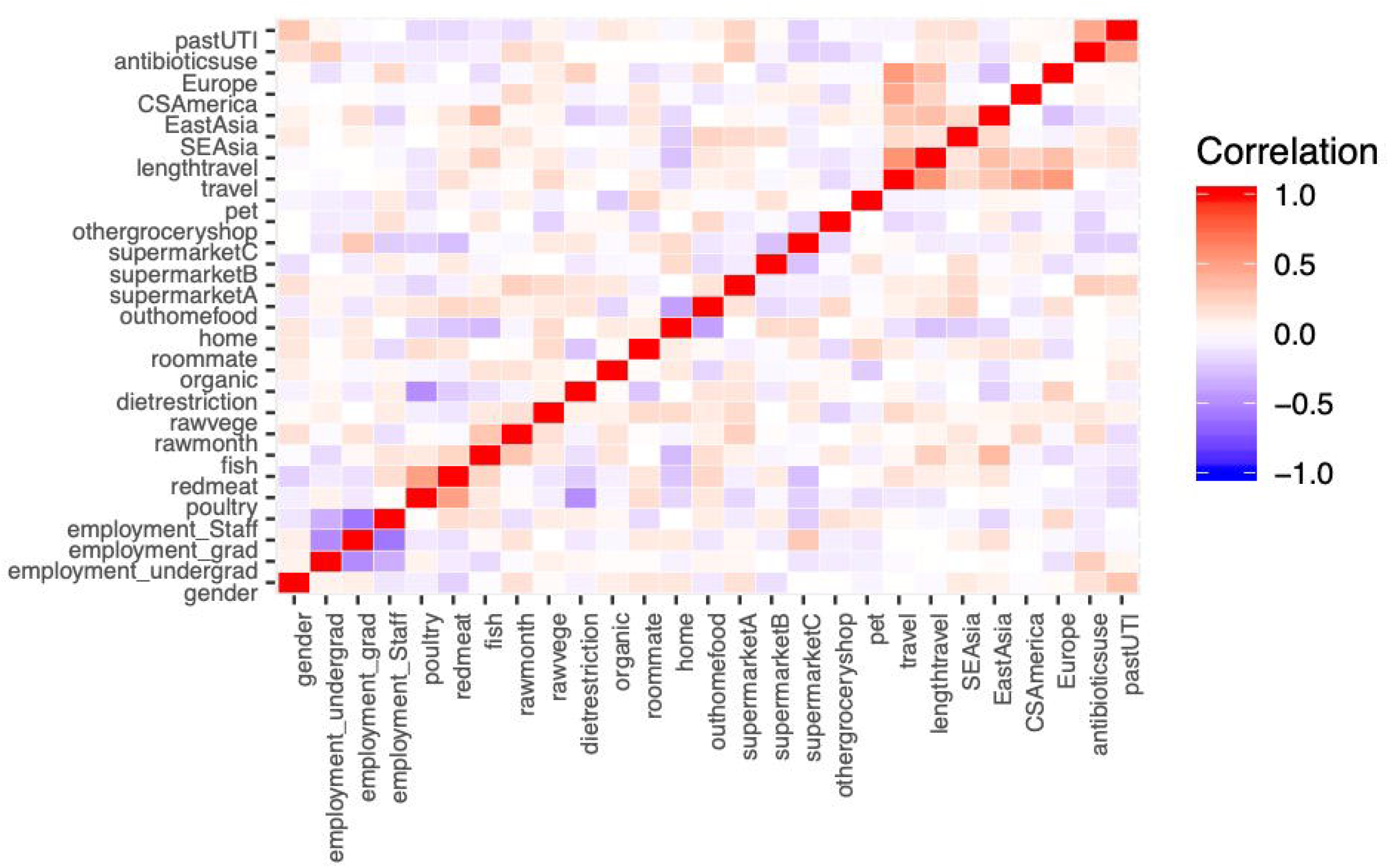
Correlation matrix for potential risk factors

### 3.2 Univariate Analysis

The risk factors associated with the fecal carriage of any drug resistant and multidrug-resistant *E. coli* based on univariate analyses are shown in Table 1. Any drug resistance was compared to those carrying drug-susceptible *E. coli*, while multidrug-resistance was compared to drug-susceptible and any drug-resistant *E. coli*. For the carriage of drug resistant *E. coli*, frequent usage of Supermarket A and previous antibiotic use had a significant association (OR=3.4[95% confidence interval (CI95%)1.1-12.4], 4.6 [1.1-31], respectively). All participants who reported to have had past urinary tract infection (UTI) carried drug resistant *E. coli*. Female gender, undergraduate status, and red meat consumption were significantly associated with carriage of multidrug-resistant *E. coli* (OR = 2.8 [1.1-7.2], 3.9 [1.3-13.2], 2.6 [1.0-6.8], respectively).

### 3.3 Multivariate Analysis

The risk factors associated with the fecal carriage of drug and multidrug-resistant *E. coli* were assessed based on multivariate logistic model (Table 2, Supplementary Table 2). The best fitting multivariate logistic model was determined with a backward elimination method based on AIC.

**Table 2:** Result of multivariate logistic regression with backward elimination model and LASSO model

|  | Multivariate logistic regression |  |  | LASSO<br>regression |
| --- | --- | --- | --- | --- |
|  | OR | 95%CI | P-value | OR |
| Female gender | 6.06 | 1.92-22.5 | 0.004 | 1.75 |
| Undergraduate | 5.46 | 1.47-25.1 | 0.017 | 1.96 |
| Red meat | 6.13 | 1.83-24.2 | 0.005 | 1.82 |
| Fish | 0.27 | 0.08-0.85 | 0.032 | 0.81 |
| Supermarket B | 2.56 | 0.87-8.28 | 0.098 | 1.07 |
| Travel within 1 year | 2.29 | 0.87-8.28 | 0.14 | - |
| Travel to Europe | - | - | - | 1.12 |
| AUC | 0.82 |  |  | 0.79 |

Fish consumption was negatively associated with the carriage of drug resistant *E. coli*, (OR = 0.17 [0.03-0.78]) whereas frequent usage of Supermarket A was a risk factor for drug-resistant *E. coli* colonization (OR = 4.5 [1.1-23]). Female gender (OR=2.3 [0.76-7.4]), organic food consumption(OR = 5.7 [0.67-150]), frequent usage of Supermarket B (OR = 3.5 [0.76-22]), and previous antibiotics use (OR = 4.3 [0.89-32]) included in the model showed positive association with the carriage of drug resistant *E. coli*, but the association was not significant when adjusted for other variables included in the model.

The variables significantly associated with the carriage of multidrug-resistant *E. coli* include female gender (OR=6.1 [1.9-22]), being undergraduate student (OR=5.5 [1.5-25]), and frequent red meat consumption (OR=6.1 [1.8-24]). Frequent fish consumption was negatively associated with multidrug-resistant *E. coli* carriage (OR = 0.27 [0.08-0.85]). Frequent usage of Supermarket B (OR = 2.6 [0.87-8.3]) and previous travel (OR=2.3 [0.76-7.4]) included in the model showed positive association with the carriage of multidrug-resistant *E. coli*, but the association was not significant when adjusted for other variables included in the model.

### 3.4 Penalized regression with LASSO

We performed penalized regression with LASSO for parameter selection. Ten-fold cross-validation was conducted to select optimal estimators. At sigma = 0.05823413, the mean squared error (MSE) showed the minimum and therefore we selected coefficients at sigma = 0.05823413 as optimum estimators (Figure 3). Estimators are shown in Table 2. Parameters included were female gender (OR=1.75), undergraduate student (OR = 1.96), frequent red meat consumption (OR = 1.82), frequent fish consumption (OR = 0.81), frequent usage of Supermarket B (OR = 1.07), and previous travel to Europe (OR = 1.12). Of these, all parameters other than previous travel to Europe were consistent with the results obtained from the multivariate model selection based on AIC.

**Figure 3:**
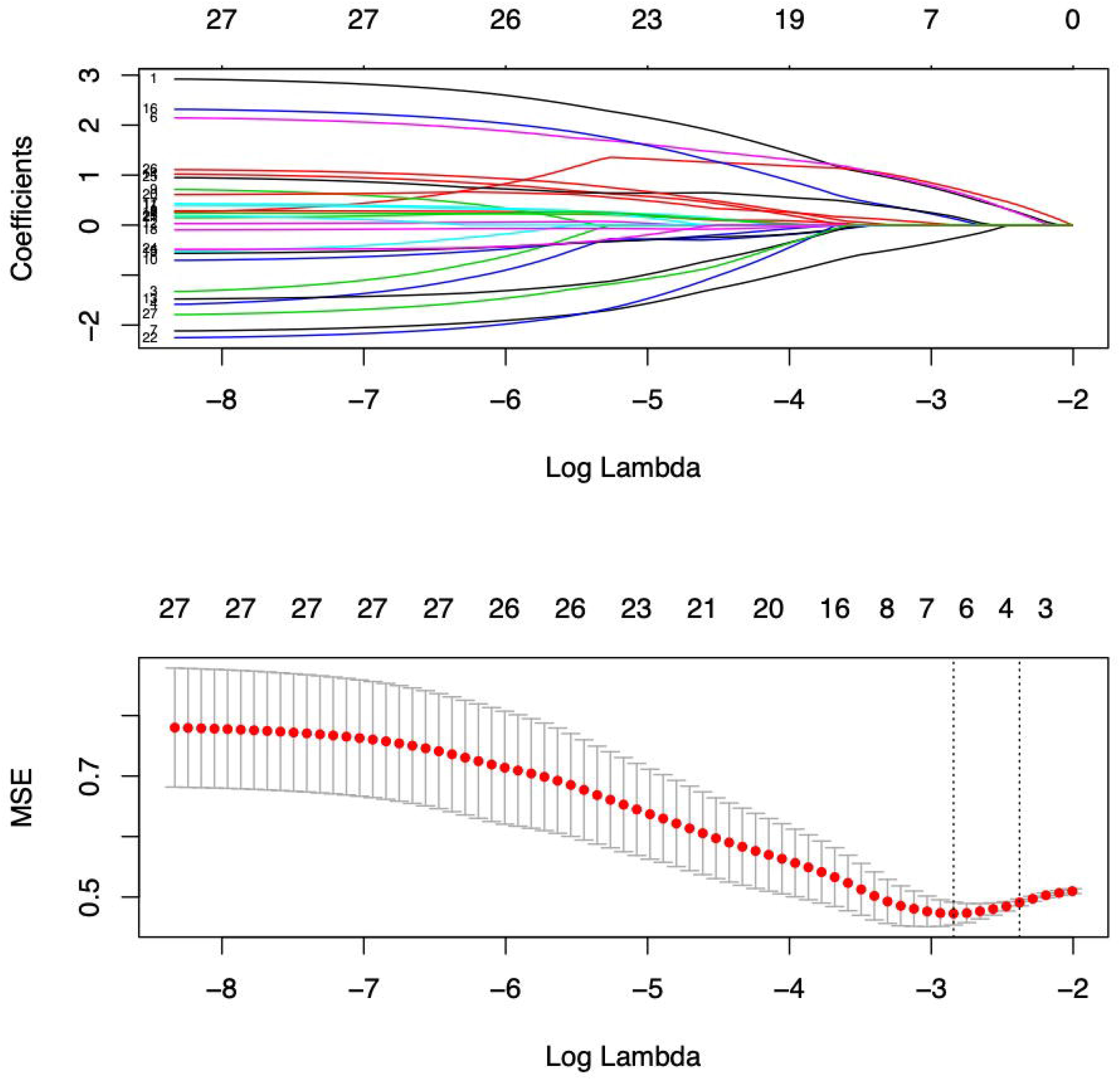
Plots for LASSO regression coefficients and cross validation over different values of the penalty parameter MSE: mean squared error

### 3.5 Model comparison

The model performance was evaluated by receiver operating characteristic (ROC) curves (Figure 4). The area under the curves (AUC) of LASSO model and backward elimination method model were 0.79 and 0.82, respectively (p=0.41). There was no difference in discrimination accuracy between LASSO model and the model with a backward elimination method.

**Figure 4:**
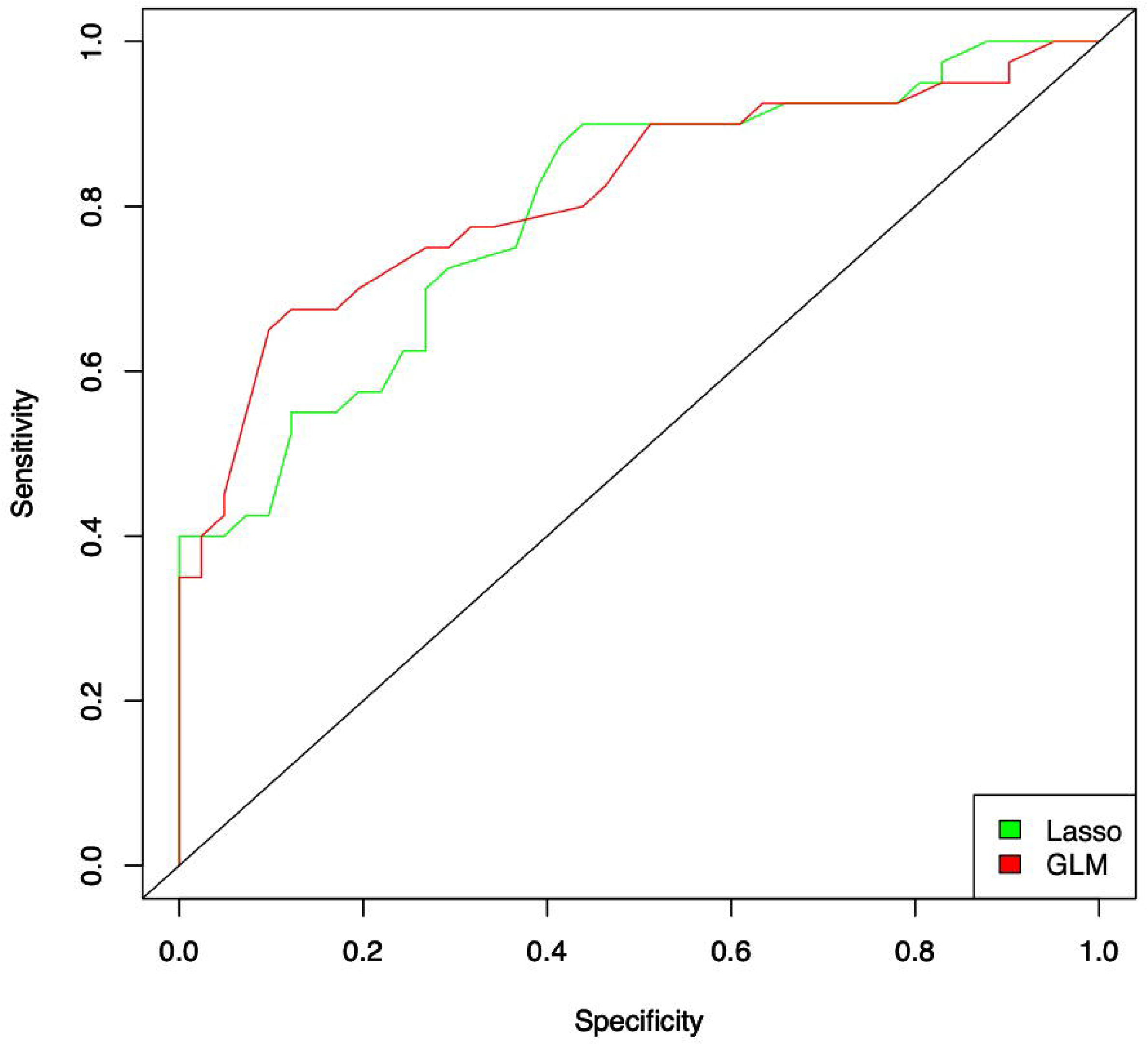
ROC curve Discrimination performance of LASSO model for fecal carriage of multidrug-resistant *E. coli* and comparison with backward elimination method model by receiver-operator characteristic analyses. Red: GLM backward elimination method model, green: LASSO model.

## 4. Discussions

This study identified potential risk factors associated with fecal carriage of multidrug-resistant *E. coli* among healthy adults at a northern California college community. As far as we know, this is the first study to find association of dietary habits with carriage of multidrug-resistant *E. coli* in healthy United States adult population. Previously, our fecal sample collection and surveillance conducted at the same college community from June to October 2018 identified the prevalence of drug resistant Gram-negative bacteria among healthy volunteers and ARGs carried by these fecal bacteria [23]. We reported the high prevalence of clinically common ARGs and integrons harbored by these Gram-negative bacteria [23]. In the present study, we aimed to identify risk factors for carriage of commensal drug-resistant *E. coli*.

Overall, the prevalence of drug resistant *E. coli* was very high in our population, with 78 (84%) of 93 fecal samples showing resistance to at least one of the antimicrobial drugs tested, which included ampicillin, gentamicin, trimethoprim-sulfamethoxazole, and colistin. Of these, 48 (52%) showed resistance to at least two of these drugs. The prevalence of both drug resistant and multidrug-resistant bacteria was higher than the pooled prevalence (14%) of a previous study that showed the pooled prevalence of drug resistant *E. coli* in the world [24] and even higher than the study with highest prevalence (51%) that targeted population after international travel conducted in Germany [33]. Unadjusted ORs revealed that while carriage of drug resistant *E. coli* was positively associated with frequent usage of supermarket A and previous antibiotics use, carriage of multidrug-resistant *E. coli* was positively associated with female gender and frequent red meat consumption. Other than the frequent usage of supermarket A, these findings are consistent with a previous report showing antibiotics use and female gender to be associated with the fecal carriage of drug resistant bacteria [24] and multiple reports showing the isolation of multidrug-resistant *E. coli* from retail meat [27,34,35].

We used multiple methods to identify risks. Since our sample size was small and was subject to overfitting under multivariate logistic regression model, we used a penalized regression method LASSO as well to select features by shrinking less important coefficients to zero and to avoid overfitting [30]. Still, features selected by LASSO are highly biased and unstable. Therefore, we implemented two methods to confirm the consistency of the outcome of model selection.

Model selection for estimating the risk of multidrug-resistant *E. coli* carriage showed consistent result with univariate analysis in both multivariate logistic regression backward elimination method and LASSO regression method. Backward elimination method with AIC and LASSO method both identified female gender, being an undergraduate student, frequent red meat consumption, and frequent usage of supermarket B as positively associated and frequent fish consumption as negatively associated with the carriage of multidrug-resistant *E. coli*. Factors related to previous travel was included in both models but were not significant nor consistent. Red meat consumption had an almost 6-fold increase and fish consumption had an almost 4-fold decrease in risk of the carriage of multidrug-resistant *E. coli*, when adjusted by gender, employment status, frequently used supermarket, and travel status. Similar trend is observed for LASSO model, showing 80% increase and 20% decrease for red meat and fish consumption respectively, adjusted by gender, employment status, frequently used supermarket, and travel status.

Red meat has been recognized to contain *E. coli* strains harboring drug resistant genes and therefore it was not surprising that frequent consumption of red meat was strongly associated with the carriage of multidrug-resistant bacteria [11,35]. Fish, however, has also been reported to be colonized with multidrug-resistant bacteria and multiple drug resistance genes [36,37], and yet we observed a negative association with both drug resistant and multidrug-resistant *E. coli* carriage in multivariate logistic regression models. From this study we cannot draw any causal relationship between frequent fish consumption and low carriage of drug resistant bacteria because of the potential unmeasured confounders related to frequent fish consumption and to the carriage of drug resistant bacteria.

Model performance of multivariate logistic regression and LASSO regression was compared with ROC. For our study these two models’ performance was equally moderate. One of the limitations of our study is the small sample size and the high imbalance between the prevalence of carriage of single-drug resistant bacteria and that of drug susceptible bacteria. Although this strong imbalance itself suggests the serious public health impact of antimicrobial resistance in the study college community, we could not assess risk factor for the carriage of any drug resistant bacteria with the LASSO method. Still, we were able to determine risk factors associated with the carriage of multidrug-resistant bacteria based on the two regression methods. The second limitation of our study is that we contacted and recruited the study participants from a restricted community—a college community. The risk factors we found may not be generalizable to other populations elsewhere. Additional studies are needed to confirm that the risk factors we found occur in other regions of the United States or other countries. However, the strength of limiting the study population to a college community is that we can control for potential unmeasured confounders such as access to food, access to health care, and other socioeconomic status.

Our third limitation is that we relied on self-reports by the participants for an assessment of their dietary behavior. Our outcome of interest, the carriage of drug resistant bacteria, was systematically tested in the laboratory. Still, our study was a cross sectional study that simultaneously accessed the outcome and the exposures related to food. Therefore, we can assume overreporting or underreporting was non-differential and any measures of association would bias toward the null, if any.

## 5. Conclusion

Among healthy adults in a college community in Northern California, female gender, being an undergraduate student, and frequent consumption of red meat was significantly associated with increased risk of being colonized with multidrug-resistant *E. coli*, while frequent consumption of fish was negatively associated. Further studies with a larger population and at other locations will be essential for establishing the generalizability of our finding and to devise public health interventions that can decrease the colonization by drug resistant bacteria.

## Data Availability

Data available upon request.

## Acknowledgement

We would like to thank Dr. Reina Yamaji for her assistance in the study design process, as well as Dr. Clarissa Araujo Borges for her consistent support in classification of the study isolates. We would also like to thank the University instructors who were instrumental in recruitment for this study. We would also like to the University Department of Environmental Health and Safety for their assistance in ensuring safe shipment of biological specimens.

## Authors’ contributions

YH performed the statistical analysis; participated in data interpretation; prepared tables and figures; wrote and drafted the manuscript; and agreed to be accountable for all aspects of the work by ensuring that questions related to the accuracy or integrity of any part of the work were appropriately investigated and resolved. JR conceptualized and designed the survey; participated in data collection; conducted laboratory testing; drafted the manuscript. KM conducted laboratory testing; participated in data collection and extraction. LWR conceptualized and designed the survey; reviewed and revised the manuscript; and agreed to be accountable for all aspects of the work by ensuring that questions related to the accuracy or integrity of any part of the work were appropriately investigated and resolved.

All authors read and approved the final manuscript.

## Figures and Tables

Supplementary Table 1: Result from survey without removing NAs

Supplementary Table 2: Multivariate logistic regression for carriage of drug resistant *E. coli*

